# Proteomic Insights into the Molecular Correlations Between Coronary Artery Disease and Peripheral Artery Disease in the Project Baseline Health Study

**DOI:** 10.1101/2024.12.20.24319444

**Authors:** Aaliyah Tucker, Maggie Nguyen, Kalyani Kottilil, Lydia Coulter Kwee, Melissa A. Daubert, Neha Pagidipati, Pamela S. Douglas, Adrian F. Hernandez, Kenneth W. Mahaffey, Svati H. Shah, the Project Baseline Health Study Group

**Affiliations:** Duke Molecular Physiology Institute, Duke University School of Medicine, Durham, NC, USA; Division of Cardiology, Department of Medicine, Duke University School of Medicine, Durham, NC, USA; Duke Clinical Research Institute, Duke University School of Medicine, Durham, NC, USA; Stanford Center for Clinical Research, Department of Medicine, Stanford University School of Medicine, Stanford, CA, USA; Division of Cardiovascular Medicine, Stanford University School of Medicine, Stanford, CA, USA; Stanford Cardiovascular Institute, Stanford University School of Medicine, Stanford, CA, USA

## Abstract

Coronary artery disease (CAD) and peripheral artery disease (PAD) are common and dangerous conditions that are both driven by atherosclerosis. Despite sharing many major risk factors, their disease pathophysiology is not fully understood. In this study, we aimed to better distinguish common vs. unique biological pathways underlying CAD and PAD by proteomics profiling plasma samples from a subset of participants in the Project Baseline Health Study (PBHS). We identified 14 proteins associated with coronary artery calcium (CAC) score ≥100 and 39 proteins associated with the ankle brachial index (ABI) after covariate adjustment. Two proteins (complement component 7 [C7] and polymeric immunoglobulin receptor [PIGR]) were significantly associated with both CAC and ABI. Our work highlights the potential molecular pathways unique to these diseases, which could have therapeutic implications for personalizing clinical management.

---

Atherosclerotic disease can manifest in diverse vascular beds including the coronary arteries (CAD) and the peripheral vasculature (PAD). Although CAD and PAD have overlapping risk factors, there are marked inter-individual differences between them in development. Relatedly, the common vs. discordant disease pathophysiology is incompletely understood. Circulating proteomics profiling provides a view into systemic biologic features that might contribute to a given disease. While prior proteomic studies have identified circulating markers of atherosclerosis, they have not described common (present in both) versus unique (specific to one) circulating proteins for CAD vs. PAD [1]. Thus, we sought to determine common vs. unique proteins for CAD vs. PAD to better distinguish common vs. unique biological pathways underlying these diseases.

The study population consisted of participants enrolled in the Project Baseline Health Study (PBHS) [2]. The PBHS enrolled 2,502 participants who had biospecimens, bilateral arterial brachial index (ABI) and cardiac imaging (including non-contrast cardiac CT) performed at enrollment. Coronary artery calcium (CAC) was measured on cardiac CT through a core lab; the resulting Agatston score was used as a measure of CAD. The lowest ABI value measured was used. One CAC score outlier and non-compressible ABI values (defined as ABI>1.4) were excluded from the analysis (**Figure 1A**). Untargeted, semiquantitative liquid chromatography mass-spectrometry (LC-MS) was performed on enrollment plasma samples from N=969 randomly selected participants using the proteomics pipeline at Verily, Inc. All proteins were scaled to mean 0 and standard deviation 1 for analysis. After exclusions, N=943 and 928 participants were included in the ABI and CAC score analyses, respectively. CAC score was analyzed as a binary outcome (i.e. <100 vs. ≥100) and logistic regression of individual proteins used to assess the association of baseline protein levels with CAD. ABI was analyzed as a continuous variable, using a linear regression model for association of baseline protein levels with ABI. Significant proteins in univariate analyses were then tested in multivariate models adjusted for age, sex, self-reported race (Black/White/Other), body mass index, history of coronary artery disease (CAD), hypertension, dyslipidemia, systolic blood pressure (SBP), diabetes, smoking, low-density lipoprotein cholesterol, and statin use. Significance was determined by a false discovery rate (FDR) p<0.1 for univariate and nominal p<0.05 for multivariate analysis.

**Figure 1.**
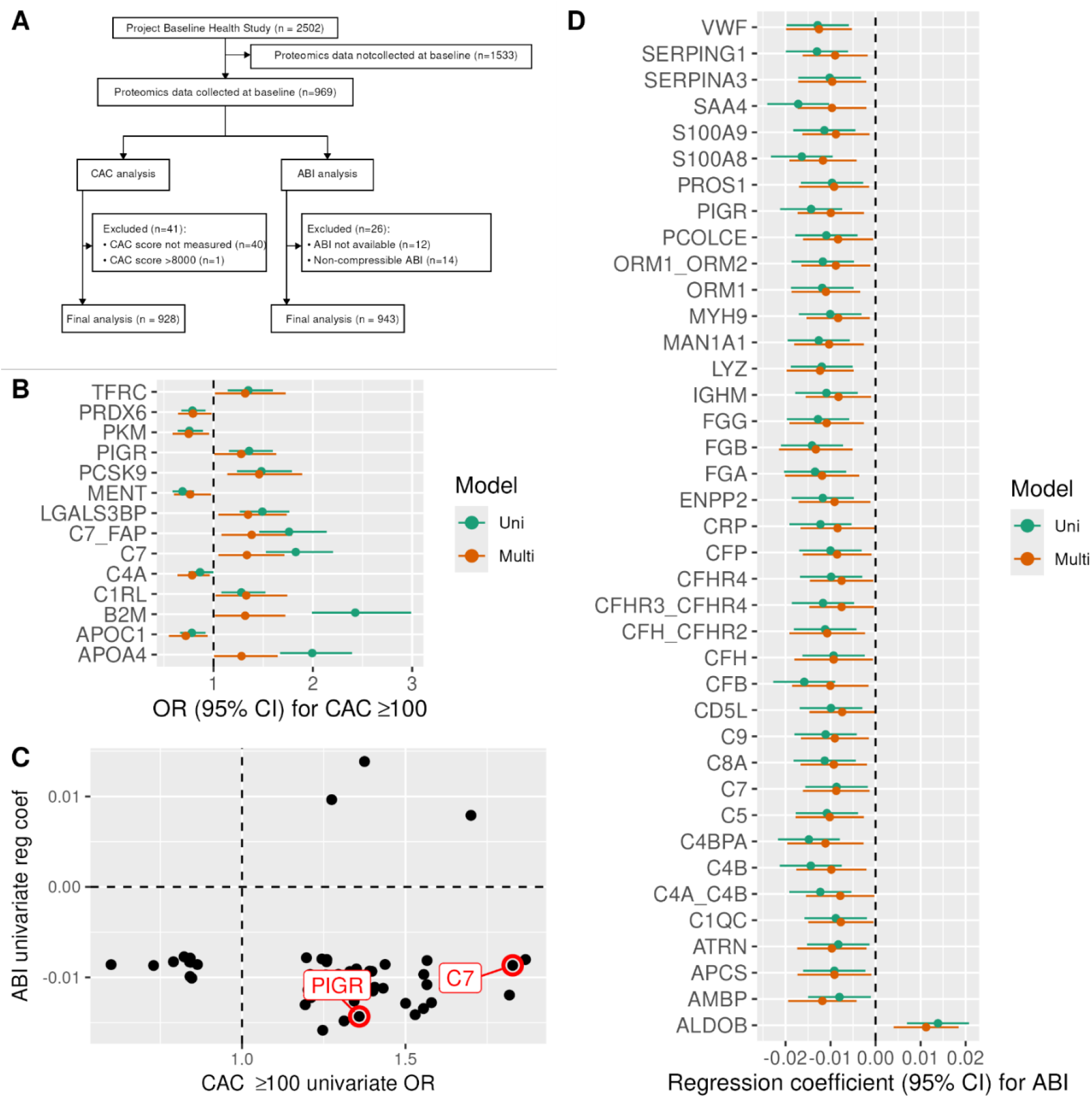
(A) Consort diagram of this study’s population. (B) Forest plot of odds ratio and 95% confidence intervals for proteins associated with CAC ≥ 100 in multivariate models. (C) Scatterplot of CAC ≥ 100 OR and ABI regression coefficients for 48 proteins significant in the univariate models. (D) Forest plot of regression coefficients and 95% confidence intervals for proteins associated with ABI in multivariate models.

Participants with CAC≥100 tended to be older, more commonly men, White, and had a history of CAD, hypertension, and diabetes, as well as higher SBP and HbA1c compared to those with CAC<100 (all p<0.001). Participants with higher ABI were also older, more likely to be men, White (all p<0.004), but not significantly different in other baseline characteristics listed above. Full baseline characteristics are included in **Supplemental Table 1**.

Of the 292 proteins profiled using mass spectrometry, 147 proteins were associated with CAC in univariate models (FDR p<0.1). After covariate adjustment, 14 proteins remained associated with CAC: lower levels of five proteins were associated with higher CAC (APOC1, C4A, PKM, MENT, and PKDX6; multivariate OR range 0.72 – 0.79, p=0.016 – 0.032) and higher levels of nine proteins were associated with higher CAC (PCSK9,C7_FAP, LGALS3BP,C7,C1RL, B2M,TFRC, PIGR, and APOA4; OR range 1.28 – 1.46, p=0.003 – 0.049) [**Figure 1B, Supplemental Table 2**]. ABI was significantly associated with 89 proteins in the univariate and 39 proteins in multivariate models; 38 of which were negatively associated with ABI (multivariate coefficient range –0.013– –0.007, p=7.7×10^-4^ – 0.046) while one (ALDOB) was positively associated (coefficient 0.011, p = 0.002) [**Figure 1D, Supplemental Table 3**]. Only two proteins (complement component 7 [C7] and polymeric immunoglobulin receptor [PIGR]) were significantly associated with both CAC and ABI after covariate adjustment **[Figure 1C]**.

Pathway analysis of proteins associated only with CAC and not with ABI using STRING (https://string-db.org/) revealed enriched pathways reporting on lipids (including “positive regulation of cholesterol esterification,” “regulation of lipoprotein lipase activity,” “high density lipoprotein particle remodeling,” “phospholipid efflux,” all FDR p<0.05). Pathway analysis of proteins associated with ABI demonstrated enrichment of pathways reporting on vascular mechanisms including hemostasis, blood coagulation, and immune response (including “fibrinolysis,” “blood coagulation,” “plasminogen activation,” “platelet activation,” “negative regulation of endothelial cell apoptotic process,” “humoral immune response,” “complement activation,” all FDR p<0.05) [3,4]. These results suggest unique molecular pathways in coronary vs. peripheral atherosclerosis, with more prominent lipid pathways for CAD vs. more immune and hemostasis pathways for peripheral disease. Surprisingly, we found only two proteins associated with both CAC and ABI. C7 is a serum glycoprotein involved in the terminal complement pathway of the innate immune system, and has been associated with unstable coronary plaque [5]. PIGR is a member of the immunoglobulin superfamily that is involved in transcytosis of immune complexes.

Thus, leveraging a deeply phenotyped cohort with systematic and direct assessments, we found that the majority of proteins associated with CAD and PAD are not common to both diseases. This highlights the potential molecular pathways unique to coronary vs. peripheral atherosclerosis, which could have therapeutic implications for personalizing clinical management.

## Data Availability

The deidentified PBHS data corresponding to this study are available upon request for the purpose of examining its reproducibility. Requests are subject to approval by PBHS governance.

## ACKNOWLEDGMENTS

The authors wish to thank Project Baseline Health Study participants and study sites.

## Funding

The Baseline Health Study and this analysis were funded by Verily Life Sciences, South San Francisco, California. Kalyani Kottilil is funded by a National Institutes of Health (NIH) F31 grant [1F31HL175914-01].

## Ethics Statement

The study was approved by the Duke University and Stanford University Institutional Review Boards. Informed consent was obtained from all participants enrolled in the Project Baseline Health Study in accordance with the Declaration of Helsinki.

## Disclosure of Conflicts of Interest

All authors acknowledge institutional research grants from Verily Life Sciences. KM reports grants from Verily, Afferent, the American Heart Association (AHA), Cardiva Medical Inc, Gilead, Luitpold, Medtronic, Merck, Eidos, Ferring, Apple Inc, Sanifit, and St. Jude; grants and personal fees from Amgen, AstraZeneca, Bayer, CSL Behring, Johnson & Johnson, Novartis, and Sanofi; and personal fees from Anthos, Applied Therapeutics, Elsevier, Inova, Intermountain Health, Medscape, Mount Sinai, Mundi Pharma, Myokardia, Novo Nordisk, Otsuka, Portola, SmartMedics, and Theravance outside the submitted work. AH reports grants from Verily; grants and personal fees from AstraZeneca, Amgen, Bayer, Merck, and Novartis; and personal fees from Boston Scientific outside the submitted work. SHS receives research funding through a sponsored research agreement to Duke University from AstraZeneca, Lilly Inc., nference Inc., Owkin, and Verily, and holds two patents on unrelated research findings. The other authors have no conflicts of interest to disclose.

